# Impact of healthcare capacity disparities on the COVID-19 vaccination coverage in the United States

**DOI:** 10.1101/2022.06.19.22276612

**Authors:** Diego F. Cuadros, Juan D. Gutierrez, Claudia M. Moreno, Santiago Escobar, DeWolfe Miller, Godfrey Musuka, Ryosuke Omori, Phillip Coule, Neil J. MacKinnon

## Abstract

**Background:** The impact of the COVID-19 vaccination campaign in the US has been hampered by a substantial geographical heterogeneity of the vaccination coverage. Several studies have proposed vaccination hesitancy as a key driver of the vaccination uptake disparities. However, the impact of other important structural determinants such as local disparities in healthcare capacity is virtually unknown.

**Methods:** In this cross-sectional study, we conducted causal inference and geospatial analyses to estimate the impact of healthcare capacity on the vaccination coverage disparity in the US. We evaluated the causal relationship between the healthcare system capacity of 2,417 US counties and their COVID-19 vaccination rate. We also conducted geospatial analyses using spatial scan statistics to identify areas with low vaccination rates.

**Findings:** We found a positive association between the healthcare capacity of a county and vaccination uptake. We estimated that a 1% increase in the Resource-Constrained Health System Index of a county increases by 0.37% the occurrence of that county in the set of counties classified as low-vaccinated (50% vaccination rate). We also found that COVID-19 vaccinations in the US exhibit a distinct spatial structure with defined “vaccination coldspots”.

**Interpretation:** We found that the healthcare capacity of a county is an important determinant of low vaccine uptake. Our study highlights that even in high-income nations, internal disparities in healthcare capacity play an important role in the health outcomes of the nation. Therefore, strengthening the funding and infrastructure of the healthcare system, particularly in rural underserved areas, should be intensified to help vulnerable communities.

## BACKGROUND

After more than two years into the pandemic, as of June 14, 2022, COVID-19 has caused 6,311,747 deaths world-wide, and the US has reported 1,011,803 of these deaths (1). Among high-income nations, the US has the highest COVID-19 mortality rate. One of the primary reasons for this is that the US has failed to achieve vaccination levels similar to those in other developed countries. As of June 2022, only 67% of the US population has been fully vaccinated against COVID-19, and this value is low compared to most European nations (1). Although this percentage is very close to the 70% goal that the US government established, if vaccination coverage is examined at the state level, the differences are striking. Vaccination coverage in the US is geographically heterogeneous. While some areas of the US have achieved full vaccination in more than 80% of their population, other regions still lag behind with rates below 50% (2, 3). A successful long-term management of the pandemic can only be achieved if vaccination uptake is substantially increased to diminish this spatial heterogeneity. However, it is necessary first to understand the factors driving the disparities in vaccination coverage and uptake in the country.

Vaccination hesitancy has been broadly discussed as a key driver of the low vaccination uptake, especially in the US (4– 6). However, COVID-19 hesitancy in the country has been estimated to be around 20%, a percentage far below the actual percentage of unvaccinated people (5), suggesting that additional unidentified key factors are behind the low vaccination rates observed in some areas of the US. It is known that the pandemic has disproportionally affected Americans living in socially vulnerable areas (7, 8). In fact, areas with low vaccination in the US experienced the highest mortality rates during the recent Delta and Omicron waves (9). Social vulnerability arises from a combination of socio-economic factors that include limited healthcare resources and barriers to accessing these resources (10). The impact of poor healthcare capacity on the vaccination coverage is a factor that has been proposed to play a major role in low-income countries, but not in high-income ones as the US (11). However, the COVID-19 pandemic has shown that the scenario is much more complex. High-income countries with better healthcare resources have had, in fact, a higher burden from COVID-19 than low-income countries with fewer healthcare resources (12). Although the US, as a nation, ranks number one in the Global Health Security Index that measures the capacity of a country to prepare for epidemics and pandemics (https://www.ghsindex.org/country/united-states/), at the local level the landscape is different. The healthcare system in the US is characterized by substantial variation in local infrastructure and capacity, with many underserved communities lacking adequate access to healthcare (7, 13, 14). These disparities include the number of healthcare workers and number of hospitals per capita, health insurance coverage, and healthcare funding, which have influenced the spatial structure of several health problems in the US, including chronic and mental health diseases (14, 15). However, how much these healthcare capacity disparities have affected the management of the COVID-19 pandemic is not known.

In this study, we conducted causal inference and geospatial analyses to assess the impact of the local healthcare capacity on vaccination coverage disparities in the US at the county level. Understanding the impact of healthcare capacity on vaccination coverage disparities will help refine local strategies to increase vaccination coverage in areas with the highest health needs.

## METHODS

### Variables and data sources

Institutional review board approval and informed consent were not necessary for this cross-sectional study because all data were deidentified and publicly available (Common Rule 45 CFR §46). This study follows the Strengthening the Reporting of Observational Studies in Epidemiology (STROBE) reporting guideline. We used causal inference analysis to evaluate the causal relationship between a treatment and an outcome, with treatment defined as the healthcare system capacity at the county level measured by the Resource-Constrained Health System (RCHS) index, and outcome defined as a specific county with a COVID-19 vaccination rate less than or equal to 50% of its population. The RCHS index is a measure generated as one of the five measures comprising the Surgo COVID Vaccine Uptake Index (CVAC) (16). For a given county, the RCHS index incorporates healthcare workforce per capita, infrastructure, healthcare spending, and care quality indicators. A high RCHS index value indicates a weak healthcare system capacity, whereas a low value indicates a strong healthcare system of the county. Vaccine coverage was measured as the proportion of the fully vaccinated population per US county, defined as the percentage of people who have received two doses of the mRNA Pfizer-BioNTech or Moderna vaccines, or a single dose of the Janssen/Johnson Johnson vaccine. Data for cumulative full vaccination rates in the total population at a county level were obtained from the Centers for Disease Control and Prevention (CDC) COVID data tracker for the contiguous US (17). We excluded the states of Colorado, Georgia, Texas, Virginia, and West Virginia due to incomplete or unreliable vaccination data. As a result, data from 2,417 counties were included in the analysis. Counties were classified as rural or urban based on the 2013 National Center for Health Statistics (18, 19). Cumulative vaccination rates were estimated as of March 31, 2022. For the causal inference analysis, counties were aggregated using the estimated vaccination rate county median of 50% into low (50%) and high (>50%) vaccination coverage groups. Based on the literature review and on the US COVID-19 Vaccine Coverage Index, we incorporated the Social Vulnerability Index (SVI) (20) and the Healthcare Access Barriers Index (HABI) as common causes that influence the treatment and outcome in the causal analysis. Vaccine hesitancy data from the CDC at the county level (21) was included as a modifier of the outcome. A detailed description of the indexes and data sources used for this study is presented in Table 1.

| Variable | Description | Source |
| --- | --- | --- |
| Vaccination rate | Cumulative full vaccination rates in the total population at a county level | CDC COVID data tracker for the contiguous US (5) |
| Resource-Constrained Health System Index (RCHS) | This index is composed by two subthemes using the listed indicators:<br>1) Low Healthcare System Capacity<br>- Vaccination provider workforce per capita<br>- Infrastructure for vaccine administration per capita<br>2) Weak Healthcare System<br>- AHRQ Prevention quality indicator<br>- Health spending per capita<br>- Total healthcare funding | Surgo Ventures - The US COVID-19 vaccine coverage index https://surgoventures.org/resource-library/the-us-covid-19-vaccine-coverage-index-leaving-no-community-behind-in-the-covid-19-vaccine-rollout |
| Healthcare Accessibility Barriers (HABI) | This index is composed by two subthemes using the listed indicators:<br>1) Barriers due to Cost<br>- Uninsured population<br>- Proportion of adults unable to cover the cost of a doctor visit<br>2) Barriers due to Transportation<br>- Households without vehicle<br>- Transit Connectivity Index |  |
| Social Vulnerability Index (SVI) | SVI indicates the relative vulnerability of every county ranking 15 social factors, including high poverty, unemployment, education, crowded housing, minority status, and disability | https://www.atsdr.cdc.gov/placeandhealth/svi/documentation/pdf/SVI2018Documentation_01192022_1.pdf |
| Vaccine Hesitancy | Percentage of the population in each county that may be vaccine hesitant or unsure about vaccination | Assistant Secretary for Planning and Evaluation (ASPE), U.S. Census Bureau's Household Pulse Survey (HPS) https://data.cdc.gov/stories/s/Vaccine-Hesitancy-for-COVID-19/cnd2-a6zw/ |

### Causal inference analysis

Randomized controlled trials offer the most plausible unbiased estimates of the effect of a given treatment on a specific health outcome (22). However, epidemiological experiments of that type are not possible due to ethical, feasibility, or time limitations (23). Causal inference approaches using epidemiological observational data are perhaps the best alternative to estimate the causal association between a treatment and a health outcome in an epidemiological context (24) (25). In this study, we implemented a causal inference approach to assess, in an unbiased manner, the average effect of the healthcare system capacity of a US county over its COVID-19 vaccination coverage.

We designed a Directed Acyclic Graph (DAG), as illustrated in Figure 1. This non-parametric graphical model visualization represents the assumed causal relationships between the established variables of interest. We included in the DAG the SVI and the HCABI as two important confounders that could have a causal effect on both, the treatment, the RCHS index, and the outcome, low vaccination rate (50%). Another variable included in the DAG was vaccination hesitancy, which was classified as a mediator affected by the RCHS index and affecting the vaccination outcome. An unobserved variable, U, was also included in the causal analysis to account for the unmeasured variables that can be potential confounders. We used a correlation analysis to test the conditional independences assumed in the DAG in the dataset, using the package DAGitty of R version 0.3-1 (26).

**Figure 1.**
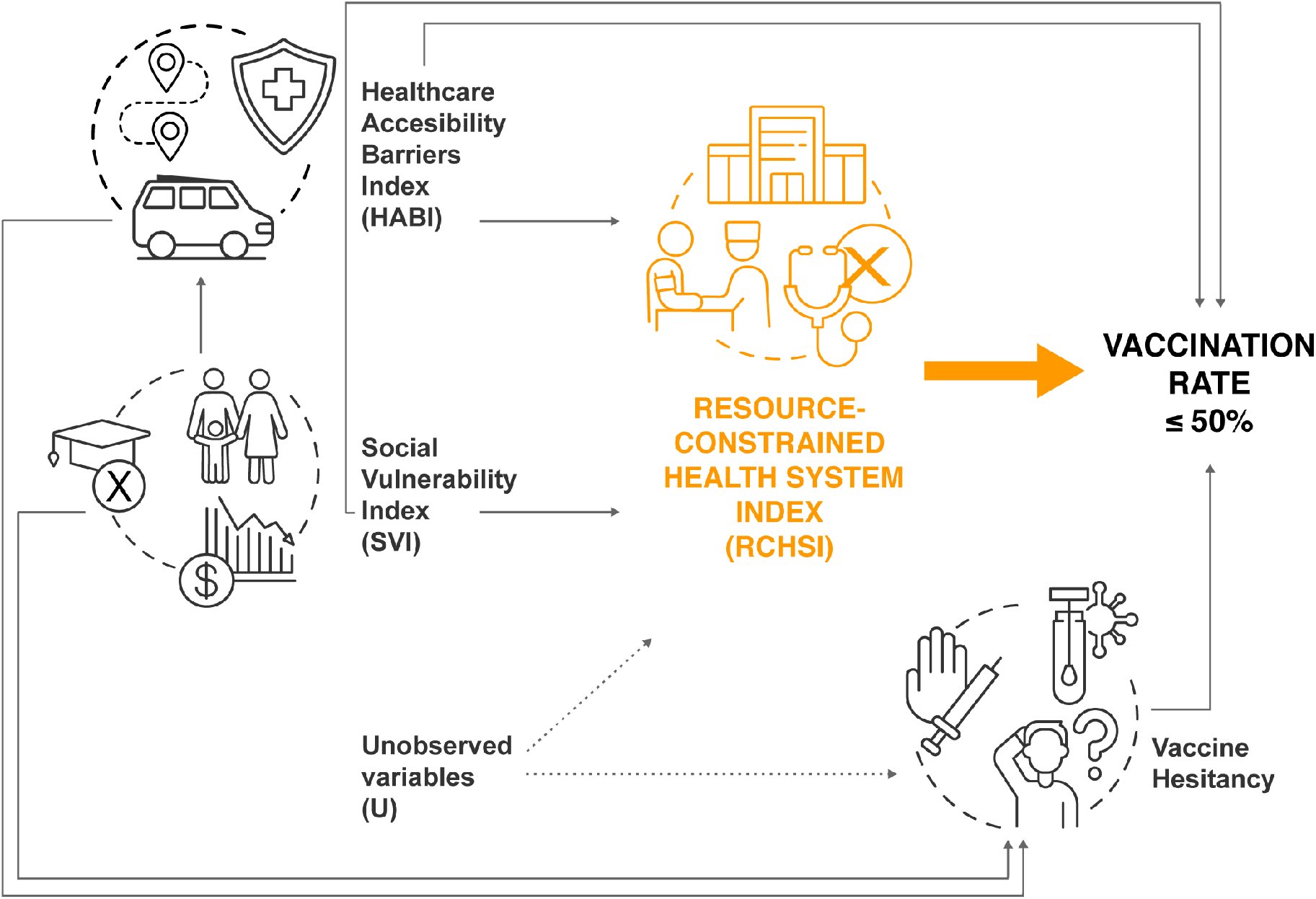
Directed Acyclic Graph (DAG) used to estimate the effect of the resource-constrained health system (RCHSI) of a county on the likelihood of this county to have a vaccination coverage 50%. The orange arrow represents the causal association of interest. Note that the variables healthcare accessibility barriers index (HABI) and social vulnerability index (SVI) are common causes that affect the treatment and the outcome simultaneously. The U variable represents the unmeasured confounder variables

**Figure 3.**
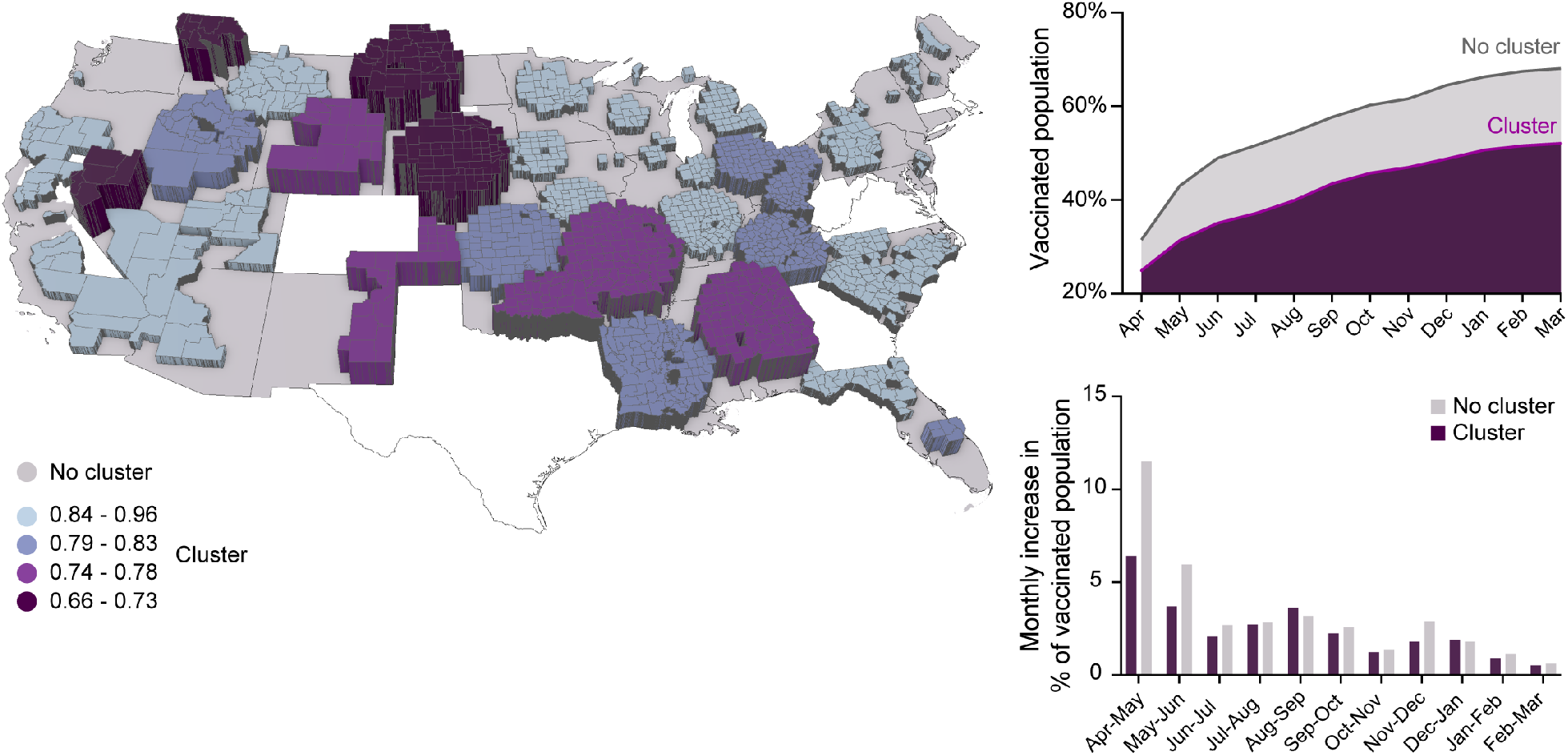
Spatial structure of COVID-19 vaccine coverage in the US. The map on the left illustrates the geospatial location of the COVID-19 vaccine coldspots. The strength of the coldspot (relative risk of being vaccinated) is illustrated with color and elevation, with the lowest relative risk for being vaccinated illustrated with dark purple, whereas the highest relative risk for being vaccinated among the coldspots being illustrated with light blue. The plot in the upper right corner illustrates the vaccination time trend within the clusters (dark purple) and outside the clusters (light grey). The plot in the lower right corner illustrates the monthly increase in percentage of vaccinated population within the clusters (dark purple bars) and outside the clusters (light gray bars) from April 2021 to March 2022.

The statistical estimand of the causal analysis, defined as the numerical value of the effect of the treatment, the RCHS index, over the occurrence of a county in the low-vaccination group was tested in a set of four variations of the double machine learning algorithm (27) using the Python modules DoWhy version 0.6 (28) and EconML version 0.13 (29). We assessed the effect of the treatment on the outcome, and low vaccination coverage at the county level using an Average Treatment Effect (ATE). Additionally, we estimated the effect of the treatment, conditioned by Hesitancy, using the Conditional Average Treatment Effect (CATE). A detailed description of the calculations and testing of the estimand can be found in the Supplementary Material. We implemented five sensitivity tests to validate the causal association between the RCHS index and low vaccination rate (50%), including the addition of a random common cause, the addition of an unobserved common cause, the replacement of a random subset, running the estimate on a random sample of the data containing measurement error in the confounders (Bootstrap refutation), and adding a placebo treatment. The dataset and the Python and R scripts used for this study are available at: https://github.com/juandavidgutier/healthcare_capacity_disparities-

### Geospatial analysis

The spatial structure of vaccination uptake was analyzed using a spatial scan statistical analysis of vaccination at the county level as of March 31, 2022, implemented in the SaTScan software (30**?**). This methodology has become the most widely used test for clustering detection in epidemiology (31), and its efficiency and accuracy are well documented (32**?**). We used scan statistics to identify geographical locations where the number of fully vaccinated individuals was lower than expected under the null hypothesis of a random spatial distribution of the vaccinated individuals across the country (30). Then, we evaluated their statistical significance by gradually scanning a circular window that spans the study region. We analyzed vaccination uptake using the SaTScan Poisson model with the size of the population at risk by location (county) included as an offset. Briefly, the identification of coldspots (areas with low vaccination rates) using the Poisson model implemented in SaTScan is achieved by testing each potential cluster against the null hypothesis that the distribution of cases (fully vaccinated individuals was proportional to the population size [no clustering] using likelihood ratio and\ t-tests) (31). An associated p-value of the statistics was then determined through Monte Carlo simulations and used to evaluate whether fully vaccinated individuals are randomly distributed in space. A coldspot was identified if the p-value was less than 0.05. After a cluster was identified, the strength of the clustering was estimated using the relative risk (RR) within the cluster versus outside the cluster. Furthermore, temporal trends of vaccination rates were analyzed by aggregating the counties within vaccination coldspots and counties outside the coldspots. Retrospective temporal vaccination rates within and outside the coldspots were estimated for each month from April 2021 to March 2022. All geographic information system (GIS) analyses and cartographic displays were performed with ArcGIS Pro version 2.9 (33) software. Plots were built using GraphPad Prism 9.

## RESULTS

As of March 31, 2022, 166,239,504 (63.1%) of 263,365,882 residents living in the counties included in the analyses were fully vaccinated. We estimated that 1,160 (48.0%) out of the 2,417 counties included in the study had a vaccination rate equal to, or lower than 50%, with 36,074,972 individuals residing in these low-vaccination counties. The average HABI was 0.54 in lower-vaccinated and 0.39 in higher-vaccinated counties. Likewise, SVI was 0.51 in lower vaccinated counties, while the average SVI in higher vaccinated areas was 0.42. Moreover, the average number of medical doctors per 10,000 in lower vaccinated areas was 7.8 compared to 18.8 in higher vaccinated areas. Similarly, the average number of ICU beds per 100,000 in lower-vaccinated counties was 11.5 compared to 18.2 ICU beds in higher-vaccinated.

### Casual inference analysis

We found nonconditional independence between the variables used in the DAG implemented to estimate the effect of the treatment, the RCHS index, on the outcome, low vaccination coverage. The machine learning algorithm with the largest RScorer was double machine learning (Rscorer = −0.0025). The ATE of the treatment, the RCHS, on the outcome, low vaccination coverage, was 0.37 (95% CI: 0.23-0.50), indicating that an increase of 1% in the RCHS index of the county has an average effect of 0.37% on the occurrence of that county in the subset of counties with low vaccination coverage. The estimation of the CATE of the treatment on the outcome conditioned by Hesitancy showed no change (Supplementary Figure 1), indicating that Hesitancy does not modify the effect of the RCHS index on the low vaccination of a county. Further results from the causal inference analysis are summarized in Supplementary Materials.

### Geospatial analysis

SatScan identified 38 clusters with low vaccination rates (vaccination coldspots) with an RR ranging from 0.66 to 0.98. These coldspots were distributed across the entire country, comprising 1,300 out of the 2,417 counties included in the study, with 930 (71.5%) of these counties being rural, compared to 612 (54.8%) of rural counties located outside the vaccination coldspots. As of March 31, 2022, the vaccination rate within the coldspots was 52.1% compared to 68.1% outside these areas. Coldspots with the lowest RR (with a RR between 0.66 and 0.73, which means that individuals residing in these vaccination coldspots had between 27% and 34% lower risk of being vaccinated compared to the areas outside the coldspots) were located in the states of Nevada, Montana, North and South Dakota, and Nebraska, with most of them grouped in the Rocky Mountain region. Vaccination coldspots with an RR between 0.74 and 0.78 were located in Idaho and in several states located in the Gulf Coast and Lower Atlantic regions, including Oklahoma, Arkansas, Mississippi, Alabama, and New Mexico. Coldspots with an RR between 0.79 and 0.83 were in the Midwest and South regions, in the states of Kansas, Indiana, Ohio, Kentucky, Tennessee, and Louisiana (map in Figure 2).

The vaccination rate was 24.9% within the low vaccination clusters, compared to 34.5% outside the coldspots at the early stage of the vaccination rollout campaign in April 2021 (area plot in Figure 2). A slower rise in the vaccination rates within the coldspots was observed during the months of May and June 2021, with 6.4% and 3.7% increments, compared to 11.5% and 6.0% increments during the same period outside the vaccination coldspots. The percentage of the vaccinated population surpassed 50% in July 2021 in counties outside the vaccination coldspots, while the same rate was reached six months later in counties within the coldspots (bar chart in Figure 2).

## DISCUSSION

In this ecological study, we found that the constraint in the health system capacity of a county was positively associated with the geographical pattern of low-vaccination uptake in the US. After controlling for other factors like vaccine hesitancy and social vulnerability, we estimated that a 1% increase in the Resource-Constrained Health System (RCHS) index of a county increases by 0.37% the occurrence of that county in the set of low vaccinated counties (50% vaccination rate). Low-vaccination areas had an average county RCHS index of 0.5, 38% higher compared to high-vaccination areas (RCHS = 0.36). In other words, our analysis showed that low-vaccination areas in the US were characterized by having a smaller vaccination provider workforce per capita, a smaller infrastructure for vaccine administration per capita, lower preventive care, and lower healthcare funding. Likewise, these low-vaccination areas were also characterized by having a higher average SVI, and higher average Healthcare Access Barrier Index compared to high-vaccination ones. We also found that vaccination in the US exhibits a distinct spatial structure with defined clustered areas of a low percentage of the population fully vaccinated (coldspots). These coldspots were distributed mainly among 17 states (NV, MT, WY, ND, SD, NE, NM, OK, MS, AL, AR, LA, TN, KY, KS, IN, and OH). Eight of these states are within the top 15 states with the highest COVID-19 mortality per capita in the US, according to the last data reported by the CDC (17). Many of these states have also been at the epicenter of the different epidemic waves in the country (7, 9), further demonstrating the strong association between lower vaccination rates and higher COVID-19 mortality within these vulnerable communities. Moreover, all of the 17 states inside the coldspots are at the bottom of a ranking evaluating healthcare access, healthcare quality, and public health in the US (34). In addition, 12 of these states fall below the US average poverty rates with 12% to 20% of their population living in poverty (35). We also found that more than 71% of the counties inside coldspots were rural.

With COVID-19 incidence and mortality increasing throughout 2020, the beginning of the immunization campaign faced unprecedented challenges that went beyond those of standard vaccination programs. Our analysis shows a clear difference between the vaccination rates in those counties that, one year later, would become vaccination coldspots. Strikingly, the rate at which vaccination uptake increased within these coldspots was much slower than the rate in the counties outside of these low-vaccinated areas, particularly at the early state of the vaccination rollout. Whereas the percentage of the vaccinated population outside the vaccination coldspots increased from 31.5% in April 2021 to 43.0% in May and reached more than 50% of the vaccination rate by July 2021, the vaccinated population within the coldspots was only 25% in April 2021, increased to 31.4% in May, and reached more than 50% vaccination rate only by January 2022. The slower vaccination uptake inside the coldspots was evident during the first three months of the period analyzed. It was relatively similar both outside of, and within the coldspots after July 2021. This suggests that pre-existent barriers in these coldspots counties played, from the beginning, an essential role in limiting the number of people who were vaccinated. Our results showed that counties inside the coldspots face a more resource-constrained health system, suggesting that critical healthcare capacity and infrastructure, and barriers to access to adequate healthcare were essential determinants of vaccination uptake. The influence of these determinants was strongly relevant during the early stages of the vaccination campaigns, a period in which vaccination availability, distribution, and prioritization needed a strong healthcare structure to deliver the maximum number of doses in the shortest time.

Collectively, these findings show the economic and health vulnerability of primarily rural communities residing in the low-vaccinated areas in the US. Rural communities within the states identified in this study may be facing challenges that exacerbate the lower rates of COVID-19 vaccination. These challenges include but might not be limited to, restricted access to testing, vaccine and treatment supplies, and number of healthcare workers(36). Interestingly, the COVID-19 vaccine coldspots were located within different geographical areas of the country, with the coldspots with the lowest vaccination rates (<45%) located mainly inside the Rocky Mountain region, coldspots with vaccination rates between 45% and 48% located in Gulf Coast and lower Atlantic regions, and coldspots with vaccination rates between 48% and 50% located in the Midwest region. Local geography can also impose challenges in the deployment of vaccines and supplies and residents’ access to rural clinics (7). Thus, it is necessary to assess if there are infrastructural disparities inside these geographical regions that can be mitigated to reduce the vulnerability of these areas.

Our study had limitations worth noting. An ecological study like the one presented here is an approach for examining the association between factors and diseases, performing population analyses in specific areas. Therefore, it is difficult to adjust for all potential confounding factors due to the lack of individual data in ecological studies. Moreover, vaccination coverage was estimated using the definition of fully vaccinated individuals, and we did not include data for boosted vaccination.

Being one of the wealthiest nations in the world, it could be assumed that the capacity of the US healthcare system is not a limiting factor in shaping national heterogeneous vaccination coverage. Hence, social and behavioral factors, such as vaccine hesitancy, have received more attention to explaining the vaccination disparities in the US. However, our study highlights that even in wealthy nations, disparities in healthcare capacity are internal determinants that play a key role in the health outcomes of the whole country. Our causal and geographical analyses unveiled a striking association between the disparities in the healthcare system capacity and the disparities of COVID-19 vaccination coverage. COVID-19 vaccines have proven to be the most effective intervention to reduce SARS-CoV-2 transmission, severity, and death. Therefore, public information campaigns and vaccine promotions, along with the strength of the healthcare system in rural underserved areas, should be intensified to target vulnerable populations in underserved communities.

Now that SARS-CoV-2 is projected to become endemic, the control of the surge of potentially dangerous new variants and seasonal epidemic outbreaks depends on the design of effective long-term immunization programs. It is key that federal, state, and county decision-makers consider the importance of strengthening the healthcare structure in these vulnerable low-vaccinated areas to increase vaccination uptake and relieve the burden that the pandemic has brought to these vulnerable communities. Healthcare disparities and differential vaccination coverage may continue to influence the pandemic trajectory and delay efforts for epidemic control. In addition, the consequences of long-term COVID- 19 will become a new challenge for the local healthcare capacity, increasing the probability of long-term health disparities in these areas.

## Data Availability

All data produced are available online at https://data.cdc.gov/Vaccinations/COVID-19-Vaccinations-in-the-United-States-County/8xkx-amqh and https://vaccine.precisionforcovid.org/

## Bibliography

1. University jh. 2019 novel coronavirus covid-19 (2019-ncov) data repository by johns hopkins csse [available from: https://github.com/cssegisanddata/covid-19,.

2. Chun-Han Lo, Leonard Chiu, Anna Qian, Muhammad Zarrar Khan, Hassan A Alhassan, Axel J Duval, and Andrew T Chan. Association of primary care physicians per capita with covid-19 vaccination rates among us counties. JAMA Network Open, 5(2):e2147920– e2147920, 2022.

3. Bhavini Patel Murthy, Natalie Sterrett, Daniel Weller, Elizabeth Zell, Laura Reynolds, Robin L Toblin, Neil Murthy, Jennifer Kriss, Charles Rose, and Betsy Cadwell. Disparities in covid-19 vaccination coverage between urban and rural counties—united states, december 14, 2020–april 10, 2021. Morbidity and Mortality Weekly Report, 70(20):759, 2021.

4. Ariel Fridman, Rachel Gershon, and Ayelet Gneezy. Covid-19 and vaccine hesitancy: A longitudinal study. PloS one, 16(4):e0250123, 2021. ISSN 1932-6203.

5. Jagdish Khubchandani, Sushil Sharma, James H Price, Michael J Wiblishauser, Manoj Sharma, and Fern J Webb. Covid-19 vaccination hesitancy in the united states: a rapid national assessment. Journal of Community Health, 46(2):270–277, 2021. ISSN 1573-3610.

6. Daniela Olivera Mesa, Alexandra B. Hogan, Oliver J. Watson, Giovanni D. Charles, Katharina Hauck, Azra C. Ghani, and Peter Winskill. Modelling the impact of vaccine hesitancy in prolonging the need for non-pharmaceutical interventions to control the covid-19 pandemic. Communications Medicine, 2(1):14, 2022. ISSN 2730-664X. doi: 10.1038/s43856-022-00075-x.

7. Diego F Cuadros, Adam J Branscum, Zindoga Mukandavire, F DeWolfe Miller, and Neil MacKinnon. Dynamics of the covid-19 epidemic in urban and rural areas in the united states. Annals of epidemiology, 59:16–20, 2021. ISSN 1047-2797.

8. Ibraheem M Karaye and Jennifer A Horney. The impact of social vulnerability on covid-19 in the us: an analysis of spatially varying relationships. American journal of preventive medicine, 59(3):317–325, 2020. ISSN 0749-3797.

9. Diego F Cuadros, F DeWolfe Miller, Susanne Awad, Philip Coule, and Neil J MacKinnon. Analysis of vaccination rates and new covid-19 infections by us county, july-august 2021. JAMA Network Open, 5(2):e2147915–e2147915, 2022.

10. J Tolbert, K Orgera, R Garfield, J Kates, and S Artiga. Vaccination is local: Covid-19 vaccination rates vary by county and key characteristics. KFF, 2021.

11. Lukman Lawal, Munira Aminu Bello, Tonderai Murwira, Clement Avoka, Shamsuddeen Yusuf Ma’aruf, Imoetin Harrison Omonhinmin, Pamela Maluleke, Christos Tsagkaris, and Helen Onyeaka. Low coverage of covid-19 vaccines in africa: current evidence and the way forward. Human Vaccines Immunotherapeutics, 18(1):2034457, 2022. ISSN 2164-5515.

12. Gabriel Demombynes. Covid-19 age-mortality curves are flatter in developing countries. 2020.

13. Ian F Miller, Alexander D Becker, Bryan T Grenfell, and C Jessica E Metcalf. Disease and healthcare burden of covid-19 in the united states. Nature Medicine, 26(8):1212–1217, 2020. ISSN 1546-170X.

14. Allison A Vanderbilt, Kim T Isringhausen, Lynn M VanderWielen, Marcie S Wright, Lyubov D Slashcheva, and Molly A Madden. Health disparities among highly vulnerable populations in the united states: a call to action for medical and oral health care. Medical education online, 18(1):20644, 2013. ISSN 1087-2981.

15. Benjamin Lê Cook, Sherry Shu-Yeu Hou, Su Yeon Lee-Tauler, Ana Maria Progovac, Frank Samson, and Maria Jose Sanchez. A review of mental health and mental health care disparities research: 2011-2014. Medical Care Research and Review, 76(6):683–710, 2019. ISSN 1077-5587.

16. Ventures s. the u.s. covid-19 vaccine coverageindex: Leaving no community behind in the covid-19 vaccine rollout. 2021 [available from:https://surgoventures.org/resource-library/the-us-covid-19-vaccine-coverage-index-leaving-no-community-behind-in-the-covid-19-vaccine-rollout,.

17. Cdc. covid data tracker 2021 [available from:https://covid.cdc.gov/covid-data-tracker/datatracker-home,.

18. ESRI. 2019 usa population density. 2020.

19. Deborah D Ingram and Sheila J Franco. 2013 nchs urban-rural classification scheme for counties. US Department of Health and Human Services, Centers for Disease Control and …, 2014.

20. Barry E Flanagan, Elaine J Hallisey, Erica Adams, and Amy Lavery. Measuring community vulnerability to natural and anthropogenic hazards: the centers for disease control and prevention’s social vulnerability index. Journal of environmental health, 80(10):34, 2018.

21. Prevention cfdca. estimates of vaccine hesitancu for covid-19 2021 [available from: https://data.cdc.gov/stories/s/Vaccine-Hesitancy-for-COVID-19/cnd2-a6zw/.

22. Angus Deaton and Nancy Cartwright. Understanding and misunderstanding randomized controlled trials. Social Science Medicine, 210:2–21, 2018. ISSN 0277-9536.

23. Franz Porzsolt and Hartmut Kliemt. Ethical and empirical limitations of randomized controlled trials. Medizinische Klinik (Munich, Germany: 1983), 103(12):836–842, 2008. ISSN 0723-5003.

24. Austin Nichols. Causal inference with observational data. The Stata Journal, 7(4):507–541, 2007. ISSN 1536-867X.

25. Peter Craig, Paul Dieppe, Sally Macintyre, Susan Michie, Irwin Nazareth, and Mark Petticrew. Developing and evaluating complex interventions an introduction to the new medical research council guidance. Evidence–Based Public Health: Effectiveness and Efficiency, 1: 185–203, 2010.

26. Johannes Textor, Benito van der Zander, Mark S Gilthorpe, Maciej LiŚkiewicz, and George TH Ellison. Robust causal inference using directed acyclic graphs: the r package ‘dagitty’. International journal of epidemiology, 45(6):1887–1894, 2016. ISSN 0300-5771.

27. Demirer M Duflo E Hansen C Newey W et al. Chernozhukov V, Chetverikov D. Double/debiased machine learning for treatment and structural parameters, 2018.

28. Amit Sharma and Emre Kiciman. Dowhy: An end-to-end library for causal inference. arXiv preprint 2011.04216, 2020.

29. Hei M Lewis G Oka P Oprescu M et al. Battocchi K, Dillon E. Econml: A python package for ml-based heterogeneous treatment effects estimation. 2019.

30. Martin Kulldorff. A spatial scan statistic. Communications in Statistics - Theory and Methods, 26(6):1481–1496, 1997. ISSN 0361-0926. doi: 10.1080/03610929708831995.

31. Martin Kulldorff and Neville Nagarwalla. Spatial disease clusters: Detection and inference. Statistics in medicine, 14:799–810, 1995.

32. G Aamodt, SO Samuelsen, and A Skrondal. A simulation study of three methods for detecting disease clusters. International Journal of Health Geographics, 5:15, 2006.

33. ESRI. Arcgis pro.x. redlands, ca, usa: Esri. 2020.

34. U.s.news. health care rankings 2022 [available from:https://www.usnews.com/news/best-states/rankings/health-care,.

35. Review wp. poverty rate by state 2022 2022 [available from: https://worldpopulationreview.com/state-rankings/poverty-rate-by-state,.

36. Michael Forster Rothbart, Kata Karáth, and Lungelo Ndhlovu. How covid-19 has exposed the weaknesses in rural healthcare. bmj, 376, 2022. ISSN 1756-1833.

