## Supplementary Materials for "Impact of healthcare capacity disparities on the COVID-19 vaccination coverage in the United States"

### Supplementary methods

#### *Data sources*

To avoid confusion bias and to estimate the causal association between the treatment, the RCHS index, and the outcome, low vaccination coverage, we included as confounders two variables: the healthcare accessibility barrier index (HABI) and the social vulnerability index (SVI). HABI is another theme of the Surgo COVID Vaccine Uptake Index CVAC that measures the cost and transportation factors limiting healthcare accessibility. It integrates measures for delayed care seeking behavior due to cost and lack of health insurance, transportation means and transit connectivity. These measurements ultimately differentiate from RCHS considering that, while RCHS measures the supply of healthcare services, HABI measures the barriers that limit access to it.

The Centers for Disease Control and Prevention (CDC) Social Vulnerability Index (SVI) measures how a community exhibits certain social conditions, including high poverty, low percentage of vehicle access, or crowded households. These conditions, according to the CDC, may affect that community's ability to prevent human suffering and financial loss in the event of disaster. The SVI is comprised of 15 variables grouped into four different categories: Socioeconomic Status, Household Composition and Disability, Minority Status and Language, and finally, Housing Type and Transportation.

We included the vaccination hesitancy rate (Hesitancy) as a mediator that the treatment, the RCHS modifies, and modifies the outcome, lower vaccination coverage. This variable was obtained from the Assistant Secretary for Planning and Evaluation (ASPE) and was calculated based on the US Census Bureau's Household Pulse Survey (HPS) data. Estimates for more granular areas such as counties were based on the data gathered at state level, using a logistic regression to analyze predictors of vaccine hesitancy using sociodemographic and geographic information. The vaccination hesitancy rate uses ordinal values to classify counties into nominal variables for Strongly Hesitant, Hesitant and Hesitant or Unsure vaccination rates at county level.

#### *Causal inference analysis*

We assessed the effect of the treatment, the RCHS index, on the outcome, low vaccination coverage at county level, through the Average Treatment Effect (ATE) (Eq. 1). Additionally, we estimated the effect of the treatment, RCHS, conditioned by Hesitancy, using the Conditional Average Treatment Effect (CATE) (Eq. 2).

$$ATE = \frac{d}{dt} E[Y(t)] \quad \text{(Equation 1)}$$

$$CATE = \frac{d}{dt} E[Y(t)|X] \quad \text{(Equation 2)}$$

where $Y$ represents the outcome, $t$ represents the treatment and the conditioning on $X$ means that we allow the treatment effect to be different depending on the characteristics of each unit, in our case we conditioned by Hesitancy.

We designed a DAG to illustrate each causal assumption in our model explicitly. Similarly, the DAG represents prior knowledge about the relationship between the variables in our causal model. As mentioned previously, we included in our DAG the variables HABI and SVI as common causes that act as confounders of the effect of the treatment, RCHSI on the outcome, and low vaccination coverage (Figure 1). The variable Hesitancy was included in the DAG as a mediator who is affected by treatment and simultaneously affects the outcome. Still, it is affected by the confounders HABI and SVI.

Through a linear regression analysis, we tested the conditional independences assumed in the DAG on our dataset. To test the conditional independence of the DAG in the dataset, we used the package DAGitty of R version 0.3-1 (1).

To identify the causal parameters, we needed to establish three assumptions:

1. Given all used potential confounders ($W$), the potential outcome $Y_t$ was independent of the exposure $T$; i.e., $Y_t \perp T|W, \forall\, t \in T$ (No unmeasured confounding)
2. No multiple versions of the treatment, i.e., $T = t \Rightarrow Y = Y(t)$ (Consistency)
3. There was a positive probability for receiving each value of treatment $T$ within every combination of covariates (Positivity)

Under these assumptions, the statistical estimand concerning the observed data were:

$$ATE = \frac{d}{d[RCHC]} (E[Low\ vac. coverage|SVI, HAB]) \quad \text{(Equation 3)}$$

$$CATE = \frac{d}{d[RCHC]} (E[Low\ vac. coverage|SVI, HAB, Hesitant]) \quad \text{(Equation 4)}$$

We estimated the statistical estimand testing a set of four variation of the double machine learning algorithm (2). This method is debiased and orthogonal and decompose the task in two stages. The first stage breaks down conditional estimations of residuals into two prediction subtasks: one for the outcome and the other for the treatment, and the second stage is a final regression with the residuals of outcome and treatment, to obtain the estimation of the ATE and CATE (2).

The tested variations of the double machine learning algorithm were as follows: linear double machine learning, sparse linear double machine learning, double machine learning, and causal forest double machine learning. To select the appropriate algorithm, we implemented the RScorer test, which is a score based on the RLearner loss (3). The RScorer fits residual models at fit time and calculates residuals of the evaluation data in a cross-fitting manner for the outcome and treatment (4). The RScorer returns an analogue of the R-square score for regression, where a negative score, means that the machine learning model performs even worse than a constant effect model and hints at overfitting during training of the model (4). Additionally, we estimated the 95% confidence interval (CI) of the effect of the treatment. Finally, we implemented five sensitivity tests to validate the causal association estimated. More specifically, we included the next sensitivity test: add a random common cause, add an unobserved common cause, replace a random subset, run the estimate on a random sample of the data containing measurement error in the confounders, aka known as Bootstrap refutation, and add a placebo treatment.

Causal inference analysis was conducted using the Python modules DoWhy version 0.6 (5) and EconML version 0.13 (4). The dataset and the Python and R scripts used for this study are available at: https://github.com/juandavidgutier/healthcare_capacity_disparities-

**Supplementary results**

We assessed the influence of a relatively small number of observations on the value of the 95% CI for the CATE and how the 95 % CI is larger for the counties with high values of the Hesitant variable (Fig. S1).

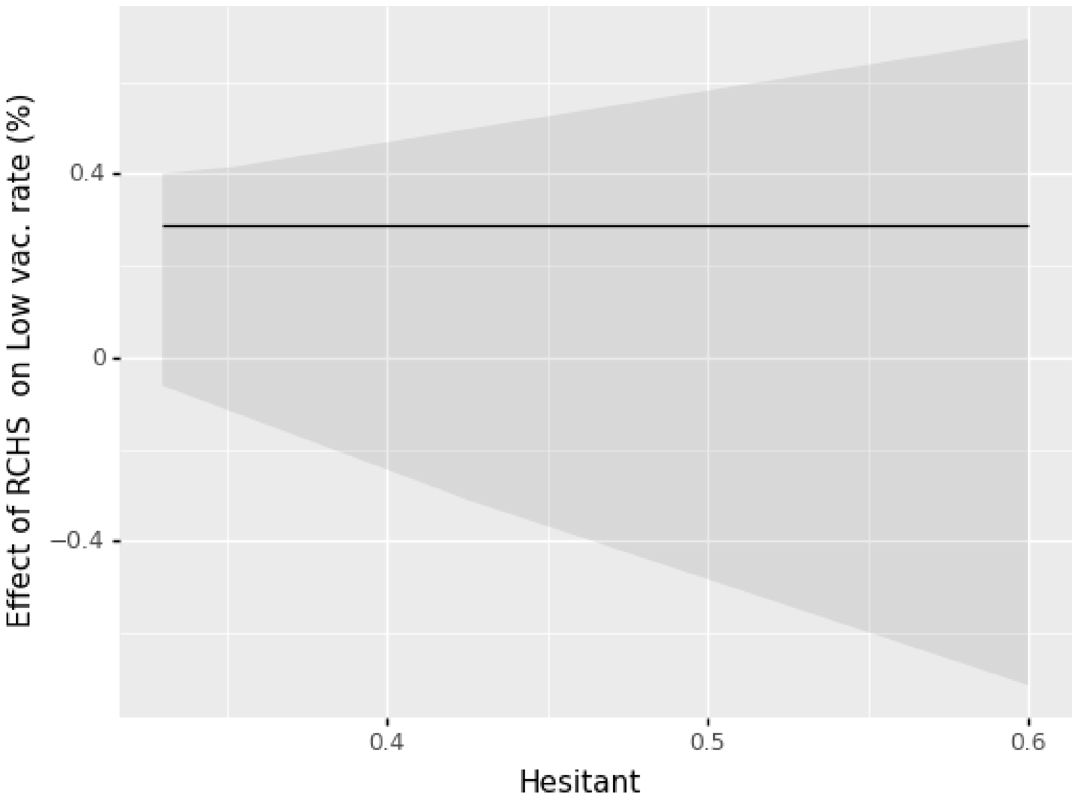


*Supplementary Figure 1. CATE and 95% CI of the effect of the treatment, the RCHS, on the outcome, low vaccination coverage, conditioned by Hesitant. The black line shows the effect of RCHS on low vaccination coverage for each value of hesitancy rate in a county (Hesitant). The grey shadow represents the 95% CI.*

The methods to test the sensibility of our model found that the values after adding a random common cause, add an unobserved common cause, replace a random subset, or include a Bootstrap refuter were close to the estimated effect (ATE) (Supplementary Table 2). The larger difference between the estimated effect and one sensibility test occurred when we implemented the Bootstrap refutation, where the difference was of 0.07 (estimated effect = 0.37 and after the Bootstrap refutation = 0.30). Similarly, the addition of a placebo treatment showed a value close to 0, which is the expected effect of a placebo treatment.

*Supplementary Table 1. Sensibility tests implemented for the effect of the treatment, RCHSI, on the outcome, low vaccination coverage. Note that for the tests of adding a random common cause, adding an unobserved common cause, replacing a random subset and Bootstrap refutation, the value should be close to the ATE. For the test of adding a placebo treatment, the value should be close to 0.*

| Estimated effect (ATE) | Add a random common cause | Add an unobserved common cause | Replace a random subset | Bootstrap refutation | Add a placebo treatment |
|---|---|---|---|---|---|
| 0.37 | 0.37 | 0.37 | 0.32 | 0.30 | 0.02 |

### References


1. Textor J, van der Zander B, Gilthorpe MS, Liśkiewicz M, Ellison GT. Robust causal inference using directed acyclic graphs: the R package 'dagitty'. International journal of epidemiology. 2016;45(6):1887-94.
2. Chernozhukov V, Chetverikov D, Demirer M, Duflo E, Hansen C, Newey W, et al. Double/debiased machine learning for treatment and structural parameters. Oxford University Press Oxford, UK; 2018.
3. Nie X, Wager S. Quasi-oracle estimation of heterogeneous treatment effects. Biometrika. 2021;108(2):299-319.
4. Battocchi K, Dillon E, Hei M, Lewis G, Oka P, Oprescu M, et al. EconML: A Python Package for ML-Based Heterogeneous Treatment Effects Estimation. 2019.
5. Sharma A, Kiciman E. DoWhy: An end-to-end library for causal inference. arXiv preprint arXiv:201104216. 2020.
